# Data management in substance use disorder treatment research: Implications from data harmonization of National Institute on Drug Abuse-funded randomized controlled trials

**DOI:** 10.1101/2020.04.28.20081935

**Authors:** Ryoko Susukida, Masoumeh Aminesmaeili, Ramin Mojtabai

## Abstract

**Background:** Secondary analysis of data from completed randomized controlled trials (RCTs) is a critical and efficient way to maximize the potential benefit from past research. De-identified primary data from completed RCTs have been increasingly available in recent years; however, the lack of standardized data products is a major barrier to further use of these valuable data. Pre-statistical harmonization of data structure, variables and codebooks across RCTs would facilitate secondary data analysis including meta-analysis and comparative effectiveness studies. We describe a data harmonization initiative to harmonize de-identified primary data from substance use disorder (SUD) treatment RCTs funded by the National Institute on Drug Abuse (NIDA) available on the NIDA Data Share website.

**Methods:** Harmonized datasets with standardized data structures, variable names, labels, and definitions and harmonized codebooks were developed for 36 completed RCTs. Common data domains were identified to bundle data files from individual RCTs according to relevant subject areas. Variables within the same instrument were harmonized if at least two RCTs used the same instrument. The structures of the harmonized data were determined based on the feedback from clinical trialists and SUD research experts.

**Results:** We have created a harmonized database of variables across 36 RCTs with a build-in label, and a brief definition for each variable. Data files from the RCTs have been consistently categorized into eight domains (enrollment, demographics, adherence, adverse events, physical health measures, mental-behavioral-cognitive health measures, self-reported substance use measures, and biologic substance use measures). Harmonized codebooks and instrument/variable concordance tables have also been developed to help identify instruments and variables of interest more easily.

**Conclusions:** The harmonized data of RCTs of SUD treatments can potentially promote future secondary data analysis of completed RCTs, allowing combining data from multiple RCTs and provide guidance for future RCTs in SUD treatment research.

## Background

Data sharing of de-identified primary data from randomized controlled trials (RCTs) has been increasingly promoted by the National Institutes of Health (NIH) (1). Secondary analysis of data from the completed RCTs is a critical and efficient way to maximize the potential benefit from past research. Designing RCTs for new treatments should ideally reflect the lessons and implications from the past RCTs. Greater availability of data would also allow investigators not involved in the primary research to confirm the findings of the original investigators, explore relationships of theoretic and clinical interest that were not examined by primary investigators, and conduct more powerful tests of hypotheses through combining multiple datasets. Furthermore, the pooled data would provide cost-effective resources for comparative efficacy studies by allowing comparisons across multiple active treatment arms. These include, for example, analyses of treatment response in under-represented population subgroups and exploration of treatment response heterogeneity in patients with risk profiles. Such data can promote new directions for research by identifying population groups who would most benefit from interventions and hence contribute to ultimately improving clinical practices.

De-identified primary data from completed RCTs have been increasingly available in recent years in various biomedical research fields. There are several organizations making critical efforts to provide researchers with user-friendly and/or harmonized RCT data for further scientific research. Some of the data repositories include the Inter-university Consortium of Political and Social Research (ICPSR), the Yale University of Open Data Access (YODA), the NIMH data archive (2), ClinicalStudyDataRequest.com (CSDR) (3), and Vivli Center for Global Clinical Research Data (4). In the field of substance use disorder (SUD) treatment, the National Institute for Drug Abuse (NIDA) has made data from the completed RCTs publicly available through the NIDA Data Share website (5) to encourage secondary analysis of completed RCTs in substance use disorder (SUD) treatment research. The NIDA Data Share website includes data from completed RCTs which were conducted under 1) the National Drug Abuse Treatment Clinical Trials Network (CTN), where patients were recruited from a broad range of community-based SUD treatment settings, and 2) the Division of Therapeutics and Medical Consequences (DTMC) program, where patients were recruited through advertisement in various academic institutions, federal therapeutic development institutions, and the pharmaceutical and biotechnology sectors to test effectiveness of innovative SUD pharmacotherapies. Deposited data from NIDA-funded RCTs are an extremely valuable resource for advancing the standard of evidence-based care for SUDs in the United States, where a high prevalence of SUDs is one of the most significant public health issues and there is a critical need for more efficacious interventions for these conditions.

As more data from RCTs have become available, there is also a growing number of secondary analyses of RCT data. However, only a limited number of secondary analyses of RCT data have been conducted and published by entirely independent investigators who were not part of the primary RCT investigative team (6). There have been increasing calls for secondary analyses of RCT data by independent investigators to ensure research reproducibility. The NIDA Data Share website has been providing a valuable resource for secondary analyses of RCT data within the SUD research community.

The NIDA CTN Dissemination Library store all publications that involved analyses of the CTN RCT data. At the time of this writing, there were a total of 636 of published documents including peer-reviewed journals, posters and newsletters, most of which were secondary analyses of data based on NIDA CTN RCTs (7). However, only 34 (5.3%) of these documents were published by entirely independent investigators, perhaps partly due to difficulty to use the RCT data from the NIDA Data Share website. This highlights the importance of maximizing the usability of the data from NIDA-funded RCTs to enable independent investigators to analyze data easily. While the primary data from completed SUD RCTs are currently downloadable from the NIDA Data Share website, the lack of standardized data products with consistent variable names, labels, and codebooks is a major barrier to further use of these valuable data. Due to these logistical data challenges, this valuable data resource is currently underutilized.

Harmonizing data structure, variables and codebooks across RCTs would facilitate secondary data analysis including meta-analysis and comparative effectiveness studies. The primary purpose of this paper is to describe a data harmonization initiative to harmonize de-identified primary data from NIDA-funded SUD treatment RCTs available on the NIDA Data Share website. We aimed to develop harmonized datasets of 36 NIDA-funded RCTs that were deposited on the NIDA Data share website by September 2018.

## Method

### NIDA-funded RCTs

There were 36 primary data publicly available from the NIDA Data Share website by September 2018, including one RCT follow-up study (CTN0030A3). We excluded open-label trials, feasibility trials, safety trials, non-randomized trials, and RCTs focusing on non-SUD outcomes from our data harmonization initiative. Table 1 presents the list of 36 RCTs included in this data harmonization initiative. There were 23 RCTs conducted under the CTN, 13 RCTs conducted under the DTMC program. By target substances, 16 RCTs targeted stimulants including cocaine and methamphetamine, 9 RCTs targeted opioids, 8 RCTs targeted any substances, and 3 RCTs targeted nicotine. By intervention types, 23 RCTs tested the effectiveness of pharmacological interventions while 13 RCTs tested the effectiveness of behavioral interventions. The sample size varied from 62 (CTN0052) to 1,285 (CTN0047).

**Table 1.** List of the studies included in the data harmonization initiative

| NIDA study code | Study title | N |
| --- | --- | --- |
| CSP999 | A multicenter clinical trial of buprenorphine in treatment of opiate dependence | 736 |
| CSP1008A | A multicenter efficacy/safety trial of buprenorphine/naloxone for the treatment of opiate dependence | 326 |
| CSP1019 | Double-blind, placebo-controlled trial of selegiline transdermal system for the treatment of cocaine dependence | 300 |
| CSP1020 | A phase III placebo-controlled, double-blind multi-site trial of lofexidine for opiate withdrawal | 68 |
| CSP1021 | Double-blind, placebo-controlled multi-center trial of baclofen for the treatment of cocaine dependence | 160 |
| CSP1022 | Phase 2, double-blind, placebo-controlled trial of selegiline transdermal system (STS) as an aid for smoking cessation | 246 |
| CSP1025 | Phase 2, double-blind, placebo-controlled trial of topiramate for the treatment of methamphetamine dependence | 140 |
| CSP1026 | Phase 2, double-blind, placebo-controlled trial of modafinil for methamphetamine dependence | 210 |
| CTN0001 | Buprenorphine/naloxone versus clonidine for inpatient opiate detoxification | 113 |
| CTN0002 | Buprenorphine/naloxone versus clonidine for outpatient opiate detoxification | 230 |
| CTN0003 | Suboxone (buprenorphine/naloxone) taper: a comparison of two schedules | 516 |
| CTN0004 | Motivational enhancement therapy (met) to improve treatment engagement and outcome in subjects seeking treatment for substance abuse | 461 |
| CTN0005 | MI (motivational interviewing) to improve treatment engagement and outcome in subjects seeking treatment for substance abuse | 423 |
| CTN0006 | Motivational incentives for enhanced drug abuse recovery: drug free clinics | 454 |
| CTN0007 | Motivational incentives for enhanced drug abuse recovery: methadone clinics | 403 |
| CTN0009 | Smoking cessation treatment with transdermal nicotine replacement therapy in substance abuse rehabilitation program | 225 |
| CTN0010 | Buprenorphine/naloxone-facilitated rehabilitation for opioid dependent adolescents/young adults | 154 |
| CTN0013 | Motivational enhancement therapy (MET) to improve treatment utilization and outcome in pregnant substance users | 200 |
| CTN0014 | Brief strategic family therapy (BSFT) for adolescent drug abusers | 482 |
| CTN0015 | Women's treatment for trauma and substance use disorders | 353 |
| CTN0021 | MET to improve treatment engagement and outcome for Spanish-speaking individuals seeking treatment for substance abuse | 436 |
| CTN0029 | A pilot study of osmotic-release methylphenidate in initiating and maintaining abstinence smokers with ADHD | 225 |
| CTN0030 | A two-phase randomized controlled clinical trial of buprenorphine/naloxone treatment plus individual drug counseling for opioid analgesic dependence | 665 |
| CTN0030A3 | Long-term follow-up to ctn-0030 (a two-phase RCTs of buprenorphine/naloxone treatment plus individual drug counseling for opioid analgesic dependence) | 375 |
| CTN0031 | Stimulant abuser groups to engage in 12-step: evaluation of a combined individual-group intervention to reduce stimulant and other drug use by increasing 12-step involvement | 471 |
| CTN0037 | Stimulant reduction intervention using dosed exercise (stride) | 302 |
| CTN0044 | Web-delivery of evidence-based, psychosocial treatment for substance use disorders | 507 |
| CTN0046 | Smoking-cessation and stimulant treatment (s-cast): evaluation of the impact of concurrent outpatient smoking-cessation and stimulant treatment on stimulant-dependence outcomes | 538 |
| CTN0047 | Screening, assessment, referral, and treatment in emergency departments (SMART-ED) | 1285 |
| CTN0048 | Cocaine use reduction with buprenorphine (curb) | 302 |
| CTN0051 | Extended-release naltrexone vs. Buprenorphine for opioid treatment | 570 |
| CTN0052 | A randomized controlled evaluation of buspirone for relapse-prevention in adults with cocaine dependence (BRAC) | 62 |
| CTO0001 | Phase 2, double-blind, placebo-controlled trial of reserpine for the treatment of cocaine dependence | 119 |
| CTO0005 | Double-blind, placebo-controlled trial of ondansetron for the treatment of cocaine dependence | 65 |
| CTO0007 | Phase 2, double-blind, placebo-controlled trial of cabergoline for the treatment of cocaine dependence | 140 |
| MDS0004 | Phase 2, double-blind, placebo-controlled trial of modafinil for the treatment of cocaine dependence | 210 |
| MDS0007 | Phase 2, double-blind, placebo-controlled trial of bupropion for methamphetamine dependence | 205 |

### Development of harmonized data format

A number of instruments were utilized in each completed RCT. Instruments included standardized tools in SUD research fields such as the Addiction Severity Index (ASI) and the Clinical Opiate Withdrawal Scale (COWS). The datasets in the harmonized database were designed so that each data file contains only one instrument and that the number of data files corresponds to the number of instruments used in each RCT. Data files have been named in a consistent manner representing each RCT identifier and instrument name. Each harmonized data file has also been provided in both a comma-separated values (CSV) file and a *Stata* (version 14; Stata Corp, College Station, TX, USA) *.dta data file.

To help users identify the instrument of interest more easily, common data domains were identified to bundle each data file according to its relevant subject area. Furthermore, variables within the same instrument were harmonized if two or more RCTs used the same instrument. The structures of the harmonized data were determined based on the feedback from the advisory board member consisting of clinical trialists and SUD research experts.

### Codebooks and supplemental documentations

For some RCTs, NIDA Data Share website provided some data along with codebooks that describe the definitions of each variable (e.g., CTN0037, CTN0044); whereas, for other studies only annotated questionnaires were provided as a guide to using the data (e.g., CTN0001, CTN0004). These data were used for identifying variables, developing uniform variable names and consistent variable and value labels. We also developed standardized codebooks for the harmonized data. The standardized codebooks were produced with the *codebook* command in *Stata*, version 14 (Stata Corp, College Station, TX, USA). Furthermore, we have developed several supplemental documents including 1) a user’s guide which documents processing notes for harmonized RCT data, 2) an instrument concordance table, which allows users to identify which instruments are comparable across different RCTs, 3) a variable concordance table, which allows users to identify which variables in each instrument are comparable across different RCTs, and 4) an assessment schedule table which tabulates assessment schedule for main outcome instruments (e.g., ASI) in each study.

## Results

Since the inception of this data harmonization initiative in September 2018, we have developed a beta version of the harmonized database of 36 NIDA-funded SUD RCTs and produced standardized codebooks as well as supplemental documentation including a user’s guide, concordance tables (for instruments and variables), and assessment schedule table.

### Harmonized data structure

Figure 1 presents the data structure of the harmonized data. Each RCT comes with a number of instruments, each of which has been stored in a separate data file. Instruments include not only standardized instruments such as ASI and COWS but also non-standardized instruments such as demographics form that varies in content across studies. The number of instruments ranged from 16 to 48 per RCT. Each instrument is provided in a separate data file and data files are named in a consistent manner. For example, the data file for ASI in CTN0001 is named “CTN0001_ASI.” The list of abbreviated names for each instrument (e.g., ASI for Addiction Severity Index) is provided in one of the supplemental documents, the instrument concordance table, which is described below.

**Figure 1.**
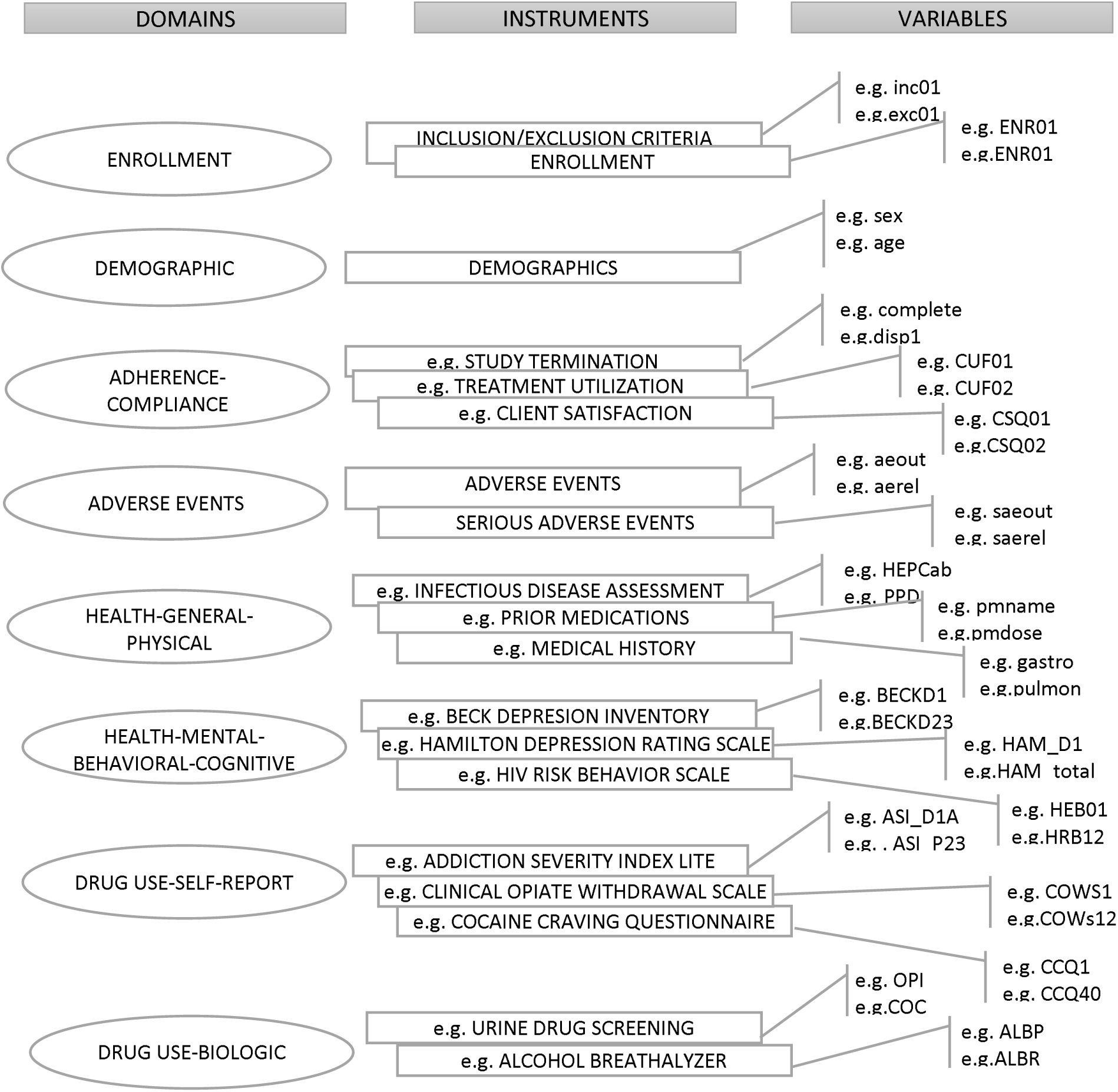
Structure of harmonized data

All the data files for each RCT have been consistently categorized into eight data domains:1) *enrollment*, containing information regarding eligibility criteria for RCT participation; 2) *demographics*, containing basic demographic information such as age, sex, and race/ethnicity; 3) *adherence*, containing information regarding the number of visits made and qualitative measures of adherence with treatments; 4) *adverse events*, containing information about medication and therapy related adverse events and often divided into severe and less than severe adverse events; 5) *physical health measures*, containing basic physical health information such as height, weight and blood pressure as well as information on medical history and prior/concomitant medications; 6) *mental-behavioral-cognitive health measures*, containing information regarding psychological well-being based on standardized tools such as Beck Depression Inventory and 36-Item Short Form Survey; 7) *self-reported substance use measures*, containing information based on self-administered tools such as ASI and Time Line Follow Back Scale; and 8) *biologic substance use measures*, containing information based on objective measures of substance use such as urine toxicology and alcohol breathalyzer.

Data presentation of variables in each data file has been made consistent across different RCTs with a build-in label and brief definition for each variable. The data files were so created that each data file starts with a set of seven variables (Figure 2) that is consistent across all datafiles. These variables allow users to combine multiple datasets from the RCT and also allow them to conduct basic analyses using each datafile separately. The first variable, “studyid”, represents the NIDA study identification code (e.g. CTN0001). The second variable, “usubjid”, represents the de-identified subject identification code. The third variable, “arm”, represents whether a patient was randomly assigned to active treatment or control arms. Names for treatment arms were taken directly from original data files. We created variables “arm1” (treatment arm for phase 1) and “arm2” (treatment arm for phase 2) for study NIDA-CTN0030/0030A, which was a two-step trial. Variable “arm” for those who were not randomized but had baseline assessments was coded either as “not randomized” or left missing. The fourth variable, “assessdays”, represents days since randomization (e.g., -3 = 3 days before randomization) for each assessment. The fourth variable, “visno” represents RCT-specific assessment visit number (e.g., WEEK4V1). The fifth variable, “phase”, represents the phase of the study, which was derived based on “visno” to allow users to distinguish whether assessments were recorded before the active treatment ("BASELINE”), during active treatment ("ACTIVE”), or after active treatment ("FOLLOWUP). Exceptionally for CTN0030/0030A, which was a two-step trial, we created two values for active phase, “ACTIVE1” (during phase 1 treatment) and “ACTIVE2” (during phase 2 treatment). The seventh variable, “measure”, represents the name of the instruments stored in a data file (e.g., ADDICTION SEVERITY INDEX-LITE). Data files for most of the measures have been structured in a long (longitudinal) format, where there are multiple assessments recorded vertically across different phases of the study. Because in many cases variables “assessdays”, “visno”, and “phase” were not all available for every assessment due to missing information in original data files, a variable “sequence” (integer starting from one) was created for some instruments, using available variables, which allows users to distinguish multiple assessments.

**Figure 2.**
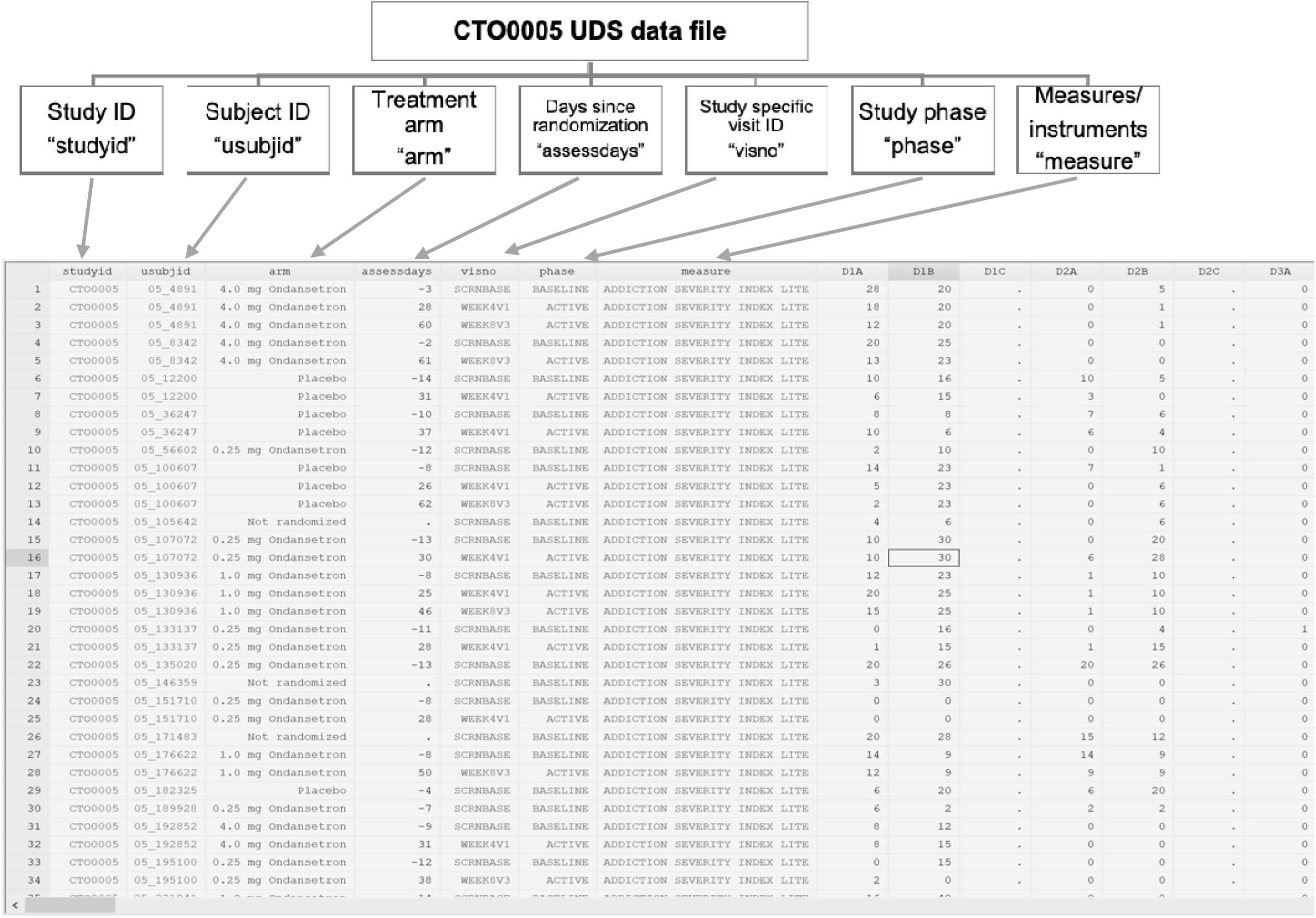
Harmonized data presentation of variables in each data file (e.g. CTO0005, Urine Drug Screening data file)

### Codebooks and supplemental documentations

Harmonized codebooks were produced using information from various documents for each CTN study available from the NIDA Data Share Website, including annotated questionnaires, protocols and spreadsheets of data dictionaries. Variable name with its brief description, variable type (string/numeric), the number of missing values, and frequency tabulation have been included for all variables. For string variables (e.g., patient id), the number of unique values has been also presented. For numeric continuous variables (e.g., quantitative measures from a urine sample), mean, standard deviation, and frequency tabulation (percentiles) have been presented. For numeric categorical variables (e.g., qualitative measures from a urine sample), range of values and variable labels for associated numeric values have been presented.

Several supplemental documents were also created. These included first, a *user’s guide*, which briefly describes how the harmonized data files were constructed, and how instruments and variables of interest can be found for secondary data analysis. The *user’s guide* also describes how data files can be merged with each other by providing several examples. Second, we have developed an *instrument concordance table* that helps users to identify which instruments/measures are included in each domain for each RCT. This document is especially useful since instruments/measures under each domain varied for every RCT (Figure 3). The *instrument concordance table* allows users to easily identify comparable instruments/measures across different RCTs. Third, we have created a *variable concordance table* since some items were omitted from instruments/measures in some RCTs (e.g., quantitative measures for urine toxicology tests were only available for a subset of 36 RCTs). The *variable concordance table* presents the item-level concordance within a particular instrument. Fourth, we have created an *assessment schedule table* showing the timing and frequencies of assessment for major SUD outcome measures such as urine toxicology tests and ASI for each RCT (Figure 4).

**Figure 3.**
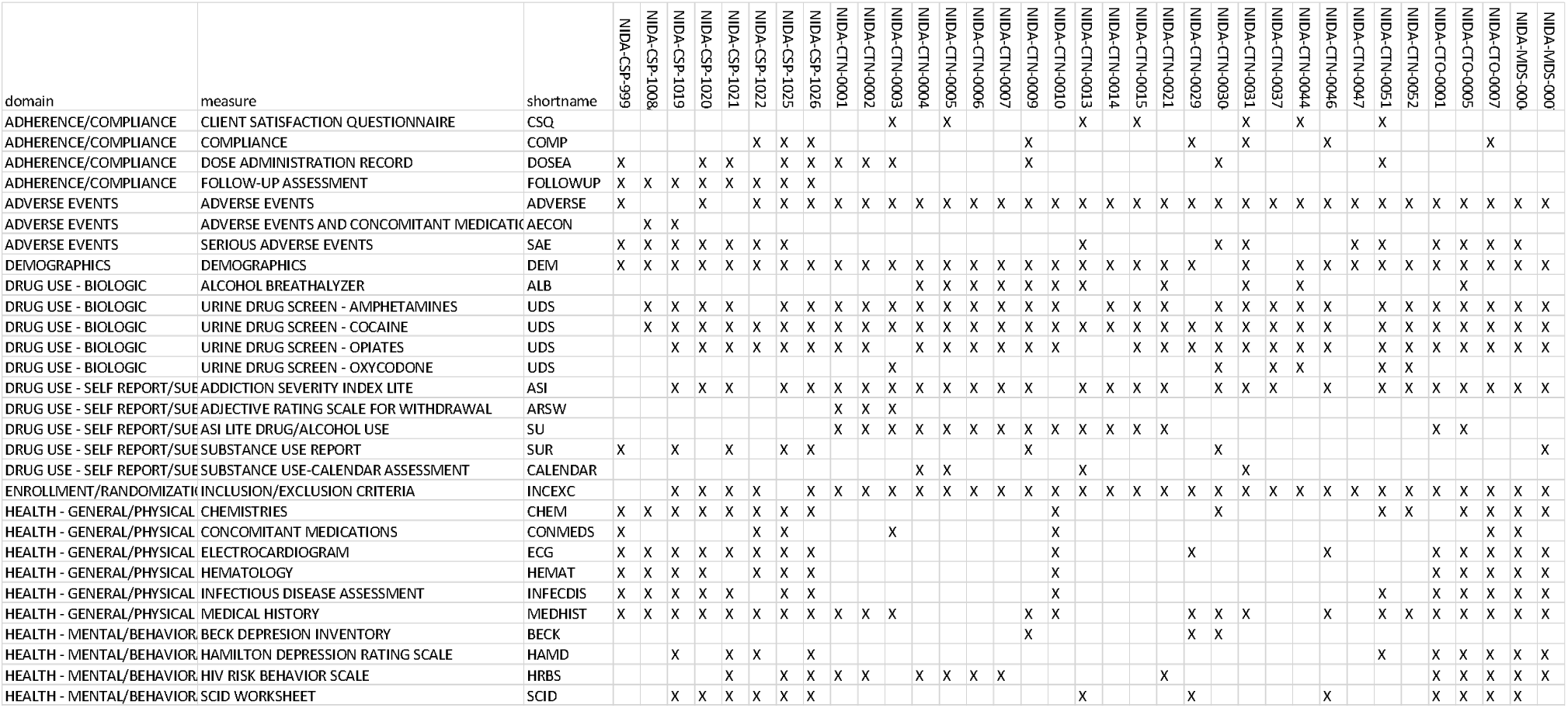
Excerpt from instrument concordance table

**Figure 4.**
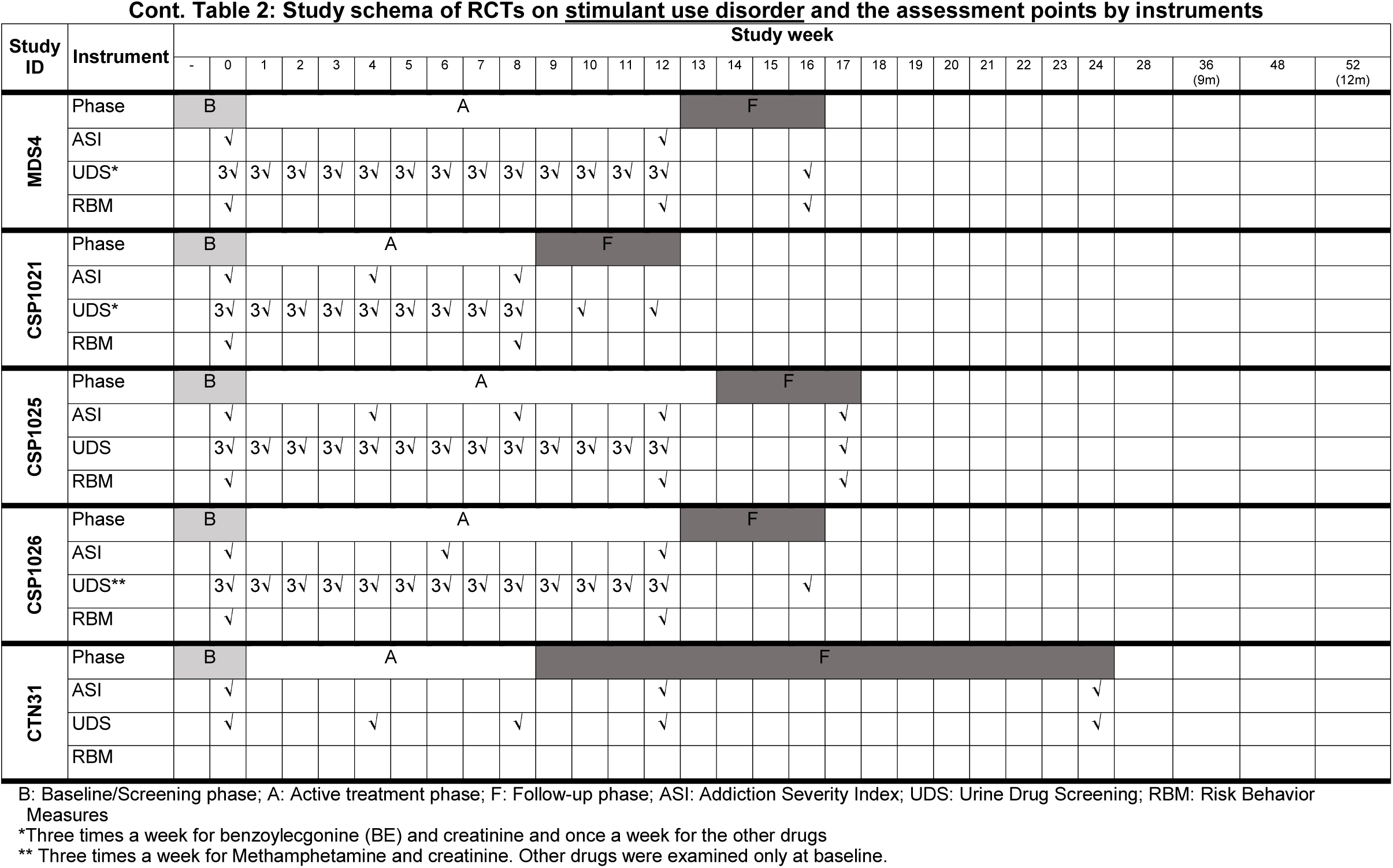
Excerpt from Assessment Schedule Table

### Examples of secondary data analyses

Table 2 presents basic descriptive statistics of 36 RCTs. Harmonized data allow researchers to easily pool data multiple RCTs and conduct comparative studies across RCTs. Secondary analyses leveraging the harmonized data have been published by our investigative team, a group of researchers who have not involved in primary data collection of the NIDA-funded RCTs (8–14). First, a series of studies by Susukida et al. (8–12) examined the generalizability of the findings from SUD RCTs to the target populations. These studies used harmonized data of NIDA-funded RCTs described above and compared the characteristics of individuals participating in SUD RCTs with individuals receiving treatment in usual care settings. The main findings of these studies were that individuals recruited into SUD RCTs appear to differ in significant was from individuals receiving treatment in usual care settings. Specifically, RCT participants had more years of education and a greater likelihood of full-time work compared with people receiving care in usual care settings. In a further step, statistical weighting was used to re-compute the effects from these SUD RCTs such that the RCT participants had characteristics that resembled those of patients in the target populations. Such re-weighting of the samples changed the treatment effects in a number of the RCTs. Most commonly, the positive effects of trials became statistically non-significant after re-weighting the RCT sample to match the target population.

**Table 2.** Socio-demographic characteristics of the harmonized data

| Study ID | Arms |  |  |  | Sample size, n | Female, % | Age, mean | White, % | Married, % |
| --- | --- | --- | --- | --- | --- | --- | --- | --- | --- |
|  | Treatment 1 | Treatment 2 | Treatment 3 | Treatment4/ placebo |  |  |  |  |  |
| CSP999 | Bup(1 mg) | Bup(4 mg) | Bup(8 mg) | Bup(16 mg) | 736 | 32.5 | 36.0 | 48.8 | 26.4 |
| CSP1008A | Bup-Nx | BU | - | Placebo | 326 | 35.0 | 37.6 | 60.4 | 22.2 |
| CSP1019 | Selegiline | - | - | Placebo | 300 | 22.0 | 40.7 | 26.5 | 18.7 |
| CSP1020 | Lofexidine | - | - | Placebo | 68 | 13.2 | 41.3 | 51.5 | 17.6 |
| CSP1021 | Baclofen | - | - | Placebo | 160 | 20.6 | 42.7 | 25.6 | 20.6 |
| CSP1022 | Selegiline | - | - | Placebo | 246 | 49.2 | 46.5 | 68.7 | 43.5 |
| CSP1025 | Topiramate | - | - | Placebo | 140 | 36.4 | 38.0 | 77.9 | 18.6 |
| CSP1026 | Modafinil 200 | Modafinil 400 | - | Placebo | 210 | 40.9 | 39.0 | 73.3 | 21.9 |
| CTN0001 | Bup-Nx | Clonidine | - | - | 113 | 39.8 | 36.2 | 55.7 | 23.0 |
| CTN0002 | Bup-Nx | Clonidine | - | - | 230 | 28.3 | 38.8 | 40.0 | 15.7 |
| CTN0003 | Bup-Nx(7-d taper) | Bup-Nx(28-d taper) | - | - | 516 | 33.0 | 35.9 | 76.5 | 24.2 |
| CTN0004 | MET | - | - | ST | 461 | 29.1 | 35.3 | 42.0 | 15.6 |
| CTN0005 | MI | - | - | ST | 423 | 42.1 | 33.3 | 71.9 | 17.0 |
| CTN0006 | Incentives | - | - | TAU | 454 | 54.9 | 35.9 | 35.7 | 18.5 |
| CTN0007 | Incentives | - | - | TAU | 403 | 43.9 | 41.9 | 25.6 | 10.9 |
| CTN0009 | SmC | - | - | TAU | 225 | 48.0 | 42.1 | 36.0 | 18.7 |
| CTN0010 | Bup-Nx(12-week) | Bup-Nx(14-day) | - | - | 154 | 41.6 | 19.7 | 70.1 | 5.2 |
| CTN0013 | MET | - | - | ST | 200 | 100.0 | 26.2 | 37.2 | 14.5 |
| CTN0014 | BFST | - | - | TAU | 482 | 21.4 | 16.0 | 40.8 | - |
| CTN0015 | SS | WHE | - | - | 353 | 100.0 | 39.2 | 48.9 | 15.9 |
| CTN0021 | MET | - | - | CAU | 436 | 10.8 | 32.7 | 0.0 | 29.4 |
| CTN0029 | ORMeth | - | - | Placebo | 255 | 43.5 | 37.8 | 80.4 | - |
| CTN0030 | Bup-Nx+SC | Bup-x+EIC | - | - | 665 | 39.6 | 33.2 | 90.8 | 28.6 |
| CTN0031 | STAGE-12 | - | - | TAU | 471 | 58.8 | 38.3 | 50.3 | 12.5 |
| CTN0037 | Exercise | Health Education | - | - | 302 | 40.1 | 38.5 | 45.4 | 13.2 |
| CTN0044 | TES | - | - | TAU | 507 | 37.9 | 35.4 | 57.6 | 14.2 |
| CTN0046 | TAU+SmC | - | - | TAU | 538 | 48.0 | 37.0 | 59.5 | 10.8 |
| CTN0047 | BS+B | SAR | MS | - | 1285 | 30.1 | 36.4 | 38.0 | 9.5 |
| CTN0051 | Nalt-ER | Bup-Nx | - | - | 570 | 33.9 | 33.9 | 69.1 | 9.1 |
| CTN0052 | Buspirone | - | - | Placebo | 62 | 37.1 | 46.1 | 25.8 | 12.9 |
| CTO0001 | Reserpine | - | - | Placebo | 119 | 29.4 | 41.0 | 21.0 | 17.8 |
| CTO0005 | Ondansetron 0.25 mg | Ondansetron 1 mg | Ondansetron 4 mg | Placebo | 65 | 15.4 | 36.2 | 38.5 | 20.0 |
| CTO0007 | Cabergoline | - | - | Placebo | 140 | 15.0 | 40.1 | 37.9 | 25.0 |
| MDS0004 | Modafinil 200 | Modafinil 400 | - | Placebo | 210 | 28.1 | 41.8 | 39.2 | 18.6 |
| MDS0007 | Bupropion | - | - | Placebo | 205 | 35.1 | 39.3 | 68.3 | 15.6 |

Abbreviations: Bup: Buprenorphine; Bup-Nx: Buprenorphine-Naloxone; BS+B: Brief Intervention plus Booster; BSFT: Brief Strategic Family Therapy; CAU: Counseling as usual; EIC: Enhanced Individual Counseling; Hisp: Hispanic; HS: High School; MET: Motivational Enhancement Therapy; MI: Motivational Interviewing; MS: Minimal Screening; NA: Not-applicable; Nalt-ER: Naltrexone-ER; NH black: Non-Hispanic black; NH white: Non-Hispanic white; ORMeth: Osmotic-Release Methylphenidate; SAR: Screening, Assessment, and Referral; SC: Standard Counseling; SmC: Smoking cessation; ST: Standard treatment; SS: Seeking Safety; TAU: Treatment as usual TES: Therapeutic Education System; WHE: Women's Health Education

Second, we conducted a study assessing the validity of the psychiatric problems subscale of the ASI (ASI-psych) (13), a detailed measure commonly used in SUD treatment research, to detect psychiatric comorbidity using pooled harmonized data from 11 NIDA-funded RCTs involving 1,660 participants. Our results demonstrated moderate accuracy of the ASI-psych in detecting the presence of any serious mental disorder against the gold standard of diagnosis based on structured or semi-structured interviews. The results support the utility of ASI-psych in screening psychiatric comorbidity among patients receiving substance use disorder treatments in RCT settings.

Furthermore, in collaboration with biostatistician, we have conducted a study to examine precision and power gains associated with adjustment for baseline variables in stratified RCTs, using the harmonized data of three stratified NIDA RCTs (14). The results demonstrated that the variance of the treatment estimates reduced up to 35% by adjusting for baseline variables, indicating that researchers planning to perform adjustment for strata and additional baseline variables could achieve the same precision with approximately 35% fewer participants.

## Conclusions

As the country struggles to tackle the growing problem of SUDs and particularly, the recent opioid epidemic, there is a pressing need for reproducible and generalizable studies to examine various interventions. The NIDA Data Share website includes data from numerous studies involving thousands of patients treated for various SUDs. Secondary analyses of these data can provide useful information by addressing new questions about treatment of SUDs and formulating new hypotheses. Our data harmonization initiative has made these data more accessible and user-friendly to a wide circle of researchers, which would encourage various secondary data analyses and help advance the standard of research and potentially ultimately, treatment, of various SUDs. The societal benefits from the deliverables from this data harmonization initiative are expected to be quite large and timely in the face of critical public health concerns in our nation. Future directions and next steps include the effective dissemination of the harmonized data to encourage secondary data analyses in the broader scientific community.

## Data Availability

The data referred to in the manuscript are publicly available from the National Institute for Drug Abuse (NIDA) Data Share website.

https://datashare.nida.nih.gov/

## Acknowledgments

We thank the advisory board members (Dr. Kathleen T. Brady; Dr. Cynthia Campbell; Dr. Walter Ling; Dr. John Rotrosen; Dr. Betty Tai; Dr. Xiaoming Wang; Dr. Li-Tzy Wu) for their valuable feedback on this data harmonization initiative.

## Declarations of competing interest

Dr. Susukida and Dr. Aminesmaeili have nothing to disclose. Dr. Mojtabai report grants from the National Institute on Drug Abuse and National Institute of Mental Health during the conduct of the study. Dr. Mojtabai has received research funding and consulting fees from Bristol-Myers Squibb and Lundbeck Pharmaceuticals.

## Notes

Funding: This project was supported by a research award from Arnold Ventures. The content is solely the responsibility of the authors and does not necessarily represent the official views of Arnold Ventures.

### Funding Statement

This project was supported by a research award from Arnold Ventures. The content is solely the responsibility of the authors and does not necessarily represent the official views of Arnold Ventures.

